# Diagnostic accuracy at the first episode of psychosis in Uganda

**DOI:** 10.1101/2020.08.28.20182501

**Authors:** Angel Nanteza, Emmanuel K. Mwesiga, Juliet Nakku, Noeline Nakasujja, Dickens Akena

**Author notes:** **Corresponding Author:** Miss Angel Nanteza, Butabika National Referral Mental Hospital.

## Abstract

**Background:** Correct clinical diagnosis at the first episode of psychosis may be difficult due to many non-specific symptoms. We aimed to determine the factors associated with a correct diagnosis among patients with a first episode of psychosis in Uganda.

**Methods:** A cross sectional study design was performed at the Butabika National Referral Mental Hospital in Uganda. We included treatment naïve participants aged 18 to 60 years with a diagnosis of a psychotic disorder. Patients with organic disorders like HIV/AIDS, syphilis and substance use disorders were excluded. The MINI international neuropsychiatric inventory was administered to confirm the clinical diagnosis. Concordance was based on the percentage agreement and kappa statistic between the admission chart diagnosis and the MINI diagnosis.

**Results:** 178 participants with a first episode of psychosis were included into the final analysis. The agreement between the MINI diagnosis and clinician diagnosis was 0.385, (P < 0.001) with a concordance of 49.5%. After controlling for nationality and the household’s source of income, duration of untreated psychosis, [p-value 0.028(95%CI: 0.07-0.89)], living with a primary family member, [p-value 0.038(95%CI:0.95-2.86)] and cadre of the clinician who made the initial diagnosis[Medical officer, [p-value 0.011(95%CI: 0.18-0.80)] were associated with a correct diagnosis.

**Conclusion:** We found low agreement between clinician diagnoses and MINI diagnoses at the first episode of psychosis. Improved training of staff while considering the duration of untreated psychosis and the living arrangements of the patient are required to improve diagnostic accuracy in this population.

## BACKGROUND

Psychotic disorders account for 10% of global burden of disease (Rathod et al., 2017; Whiteford, Ferrari, Degenhardt, Feigin, & Vos, 2015). Approximately 80% of the burden of psychotic disorders is found in low and middle income countries (LMIC) like Uganda (Kigozi et al., 2016; Organization, 2017; Rathod et al., 2017). Psychotic disorders are associated with high productivity loss and mortality due to suicide (Laursen, Munk-Olsen, & Gasse, 2011; Lencz et al., 2014; Rost, Meng, & Xu, 2014). In order to reduce the burden associated with these disorders it is essential that they are identified, diagnosed and managed early in the course of the illness (Marshall & Rathbone, 2011; McGorry, 2015). It is preferable that the management is initiated early at the first episode of psychosis(FEP) (Anderson et al., 2018; Norman & Manchanda, 2016).

FEP, operationally defined as the time of initiation of antipsychotic medication; is an important time point in the course of a psychotic illness (Breitborde, Srihari, & Woods, 2009). Literature from high income countries indicate that correct management of psychotic disorders during FEP has been associated with improved outcomes like quality of life (Heeramun-Aubeeluck et al., 2015; Marshall & Rathbone). Differences in health seeking behaviour and health systems however imply that the time for initiation of antipsychotic medication varies widely (Mossaheb et al., 2013; Murru & Carpiniello, 2018; Oosthuizen, Emsley, Keyter, Niehaus, & Koen, 2005). Moreover, a number of patients present with varieties of clinical features that make it difficult to make a conclusive diagnosis and rate severity (Abbo, Ekblad, Waako, Okello, & Musisi, 2009; Mossaheb et al., 2013). Even with this challenge it is still imperative that an attempt is made to correctly classify patients according to some set diagnostic criteria. For psychotic disorders this might be difficult as many of the symptoms continuously change as the disease progresses (Dutta et al., 2007; Heckers et al., 2013; Tandon, 2013). The presence of many non-specific signs and symptoms are at FEP may therefore impact on clinicians diagnoses (Dutta et al., 2007).

The diagnostic accuracy for psychotic disorders at FEP in LMIC is not well described. In low resource settings, severe human resource constraints and greater need to task shift implies that few patients are reviewed by psychiatrists (Swartz, Kilian, Twesigye, Attah, & Chiliza, 2014). It is unclear if there is an association between clinician’s level of training and correct diagnoses among patients with FEP. The association of patient illness factors like symptom severity, age of onset of illness and duration of untreated psychosis on correct diagnoses at FEP is also not well described. We therefore aimed to determine the diagnostic accuracy between diagnoses made by clinicians against a gold standard among patients with FEP at the country’s national referral and teaching mental hospital. We also aimed to determine whether socio-demographic, clinician and illness factors were associated with the accuracy of the diagnosis.

## METHODS

### Study site

This was a cross sectional study conducted on the four acute admission wards at Butabika National Referral Mental Hospital in Uganda. Butabika National Referral and Teaching Mental Hospital is located about 12 kilometers south east of Kampala city in Nakawa sub-county. On average, a total of about 200 patients are attended to daily in the outpatient units of the hospital (Emmanuel Kiiza Mwesiga et al., 2019). The hospital has 4 general admission wards, 2 sick wards and 2 convalescent wards. There are specialized services for addiction psychiatry, social work, forensic psychiatry, child and adolescent mental health services and psychotherapy. Each general ward has about 150 in patients who are assigned to one of five different firms. These firms are supervised by clinical teams comprised of psychiatrists, residents in training, medical officers, clinical psychologists, psychiatric social workers, psychiatric nurses and psychiatric clinical officers. These teams review the patients in their firms on a regular basis.

### Ethical approvals

Ethical approval was sought from and approved by the Uganda National Council for Science and Technology (UNCST) (#HS142ES), the School of Medicine Research and Ethics Committee (SOMREC), Makerere University (#2017-153) and institutional approval was received from Butabika hospital

### Study participants

Participants for this study were selected from an ongoing study in which we aim to determine the factors associated with cognitive impairment among patients with a first episode of psychosis. This study enrolled consenting in-patients with a psychosis diagnosis aged 18 to 60 years and who were naïve to antipsychotic medication. Participants with HIV/AIDs, substance use and Syphilis were excluded.

### Study tools

The MINI International Neuropsychiatric Inventory is a semi-structured interview guide for making DSM-5 diagnoses. It has been extensively used in our setting. Only Module D (mania), module J (alcohol use), module K (other substances) and module L (psychotic disorders) were administered during the study. Specifically, for module L an extended version of the MINI was used. This extended version has an additional 12 questions that helps a clinician to subtype a specific psychotic disorder. With the extended version, one can differentiate between schizophrenia and schizophreniform disorders, which may not be possible in the non-extended version of the MINI. The non-extended version is being used in the original study (Anne Stevenson et al., 2019).

In the primary study, participants had sociodemographic information collected using a standardized questionnaire. This questionnaire collected data on sociodemographic characteristics like age, gender, religion, source of income and marital status. It also collected data on onset of psychotic symptoms which was used to calculate the duration of untreated psychosis as well as alternative therapies used before admission.

### Study procedure

All participants who were enrolled in the primary study and agreed to take part in this study had their psychosis diagnosis confirmed using the extended version of the MINI. The extended version of the MINI was administered by a psychiatric clinical officer (AN) who had been trained. A psychiatrist (EKM) supervised the assessment process. Later, chart abstraction was performed to determine the diagnosis made at admission as well the cadre (level of training) of staff who made the diagnosis.

### Statistical methods

Data was analysed in June 2019 using StataCorp (2015). Statistical Software: Release 14. College Station, TX: StataCorp LP. Diagnoses made by the different clinician cadres in the different psychiatric teams were compared to diagnoses confirmed using the MINI. Our dependant variable was the participants whose admission diagnosis correlated with the MINI diagnosis. Patient sociodemographic, illness related (age of onset, duration of untreated psychosis and age at initial contact with mental health facility) and clinician-related factors like who made the diagnosis were our independent variables. Duration of untreated psychosis (DUP) can be defined as the time from the emergence of the first psychotic symptom to the start of antipsychotic treatment (Drancourt et al., 2013). DUP was calculated by subtracting the age at which the patient first recognised symptoms from their current age. At multi-variate analysis a level of significance of less than 0.05 was used to test for significance between different factors and a correct diagnosis on both the MINI and admission file while controlling for nationality and the household’s source of income.

## RESULTS

Our final dataset included 178 participants who were assessed using the MINI. Briefly the median age was 27 years (IOR 24-36) with 51% (92/178) of the participants being female. The commonest diagnosis at admission was Bipolar Affective disorder (28%). The proportions of different diagnoses in the chart and on the MINI are shown in table 1 below.

**Table 1.** Comparison between the diagnosis made at admission and the gold standard (MINI) diagnosis.

| Diagnosis | Dx at Admission<br>Number (%) | MINI Dx<br>Number (%) |
| --- | --- | --- |
| Alcohol psychosis | 0 (0) | 14 (7.7) |
| Bipolar Affective Disorder | 51 (28.0) | 50 (27.5) |
| Brief Psychotic disorder | 29 (15.9) | 17 (9.3) |
| Major depressive disorder | 0 (0) | 1 (0.6) |
| Depression with psychotic features | 15 (8.2) | 28 (15.4) |
| Schizophrenia | 42 (23.1) | 57 (31.3) |
| Schizoaffective | 2 (1.1) | 4 (2.2) |
| Schizophreniform | 2 (1.1) | 11 (6.0) |
| Other | 23 (12.7) | 0 (0) |
| Unknown | 18 (9.9) | 0 (0) |

Agreement: Cohen's Kappa = 0.385, $P < 0.001$ and concordance was 49.5%

Table 2 shows the bivariate associations between of concordance of the admission and MINI diagnoses with various sociodemographic characteristics. There were no associations between a correct diagnosis and age [prevalence ratio (PR) 0.99 (95% confidence interval (CI) 0.73-1.35; p value 0.984] or gender age [PR 1.08 (95% CI 0.79-1.46); p value 0.643].

**Table 2:** The factors associated with the concordance between clinical diagnosis at admission and MINI Diagnosis by trained RA.

| Variable | Number (N) | Concordance [n(%)] | Prevalence ratio | 95% CI | P-value |
| --- | --- | --- | --- | --- | --- |
| **Age (categorised at median)** |  |  |  |  |  |
| Median = 27 (IQR 24 – 36) |  |  |  |  |  |
| ≤ 27 | 93 | 45 (48.4) | 1 |  |  |
| > 27 | 85 | 41 (48.2) | 0.99 | 0.73 – 1.35 | 0.984 |
| **Gender** |  |  |  |  |  |
| Male | 86 | 40 (46.5) | 1 |  |  |
| Female | 92 | 46 (50.0) | 1.08 | 0.79 – 1.46 | 0.643 |
| **Marital status** |  |  |  |  |  |
| Single | 95 | 45 (47.4) | 1 |  |  |
| Married | 51 | 27 (52.9) | 1.12 | 0.80 – 1.56 | 0.516 |
| Divorced/Widowed | 32 | 14 (43.8) | 0.92 | 0.59 – 1.45 | 0.728 |
| **Current employment** |  |  |  |  |  |
| Student | 15 | 8 (53.3) | 1 |  |  |
| Formal employment | 17 | 8 (47.1) | 0.88 | 0.44 – 1.77 | 0.724 |
| Non-formal employment | 73 | 33 (45.2) | 0.85 | 0.49 – 1.45 | 0.547 |
| Unemployed | 73 | 37 (50.7) | 0.95 | 0.56 – 1.61 | 0.850 |
| **Nationality** |  |  |  |  |  |
| Uganda | 167 | 78 (46.7) | 1 |  |  |
| Other | 10 | 7 (70.0) | 1.50 | 0.97 – 2.32 | 0.070 |
| **Main source of income in household** |  |  |  |  |  |
| Self | 63 | 26 (41.3) |  |  |  |
| Father | 29 | 16 (55.2) | 1.34 | 0.86 – 2.08 | 0.198 |
| Mother | 21 | 12 (57.1) | 1.38 | 0.86 – 2.23 | 0.179 |
| Relative/Guardian | 49 | 22 (44.9) | 1.09 | 0.71 – 1.67 | 0.700 |
| Non-relative/Organisation | 13 | 8 (61.5) | 1.49 | 0.88 – 2.51 | 0.134 |
| **Most import household source of earning** |  |  |  |  |  |
| Subsistence farming | 51 | 25 (49.0) | 1 |  |  |
| Commercial farming | 3 | 2 (66.7) | 1.36 | 0.58 – 3.18 | 0.478 |
| Wage employment | 65 | 31 (47.7) | 0.97 | 0.67 – 1.42 | 0.887 |
| Non-agricultural enterprises | 16 | 10 (62.5) | 1.23 | 0.79 – 2.05 | 0.314 |
| Property income | 19 | 7 (36.8) | 0.75 | 0.39 – 1.45 | 0.392 |
| Organisational support | 2 | 2 (100.0) | 2.04 | 1.54 – 2.70 | <0.001 |
| Others | 19 | 7 (36.8) | 0.75 | 0.39 – 1.45 | 0.392 |
| **Current living arrangements** |  |  |  |  |  |
| Renting | 30 | 10 (33.3) | 1 |  |  |
| Owns house | 38 | 19 (50.0) | 1.50 | 0.82 – 2.73 | 0.185 |
| Living with primary family | 71 | 39 (54.9) | 1.65 | 0.95 – 2.86 | 0.075 |
| Living with other family | 29 | 13 (44.8) | 1.34 | 0.70 – 2.57 | 0.371 |
| Living with friends | 3 | 1 (33.3) | 1.00 | 0.19 – 5.38 | 1.000 |
| No housing/Living in street | 6 | 3 (50.0) | 1.50 | 0.58 – 3.88 | 0.403 |
| Others | 1 | 1 (100.0) | 3.00 | 1.81 – 4.98 | <0.001 |
| **First time presenting with symptoms?** |  |  |  |  |  |
| No | 54 | 29 (53.7) | 1 |  |  |
| Yes | 124 | 57 (46.0) | 0.86 | 0.63 – 1.17 | 0.331 |
| **First time at Medical facility?** |  |  |  |  |  |
| No | 22 | 12 (54.6) | 1 |  |  |
| Yes | 152 | 73 (48.0) | 0.88 | 0.58 – 1.34 | 0.550 |
| **Given Antipsychotic medication before?** |  |  |  |  |  |
| No | 151 | 73 (48.3) | 1 |  |  |
| Yes | 24 | 13 (54.2) | 1.12 | 0.75 – 1.68 | 0.582 |
| **Age first sought help**<br>(categorised at median; Median = 26. IQR 22 – 33) |  |  |  |  |  |
| ≤ 26 | 89 | 43 (48.3) | 1 |  |  |
| > 26 | 82 | 41 (50.0) | 1.02 | 0.76 – 1.41 | 0.826 |
| **Previously used Traditional healer** |  |  |  |  |  |
| No | 123 | 61 (49.6) | 1 |  |  |
| Yes | 53 | 24 (45.3) | 0.91 | 0.65 – 1.29 | 0.607 |
| **Previously used a spiritual healer** |  |  |  |  |  |
| No | 96 | 46 (47.9) | 1 |  |  |
| Yes | 80 | 39 (48.8) | 1.02 | 0.75 – 1.38 | 0.912 |
| **Previously used a witch doctor** |  |  |  |  |  |
| No | 143 | 68 (47.6) | 1 |  |  |
| Yes | 33 | 17 (51.5) | 1.08 | 0.75 – 1.57 | 0.675 |
| **Who made initial diagnosis** |  |  |  |  |  |
| Psychiatrist | 17 | 13 (76.5) | 1.00 |  |  |
| Medical officer | 14 | 6 (42.9) | 0.56 | 0.9 – 1.09 | 0.086 |
| Clinical officer | 136 | 65 (47.8) | 0.63 | 0.45 – 0.89 | 0.004 |
| Nurse | 11 | 2 (18.2) | 0.24 | 0.07 – 0.89 | 0.028 |
| **DUP [Mean (SD)]** | 1.30 (3.71) |  | 1.03 | 1.01 – 1.05 | 0.002 |

After controlling for nationality and household’s most import source of income the variables that remain significantly associated with a correct diagnosis are shown in table 3 below.

**Table 3:**
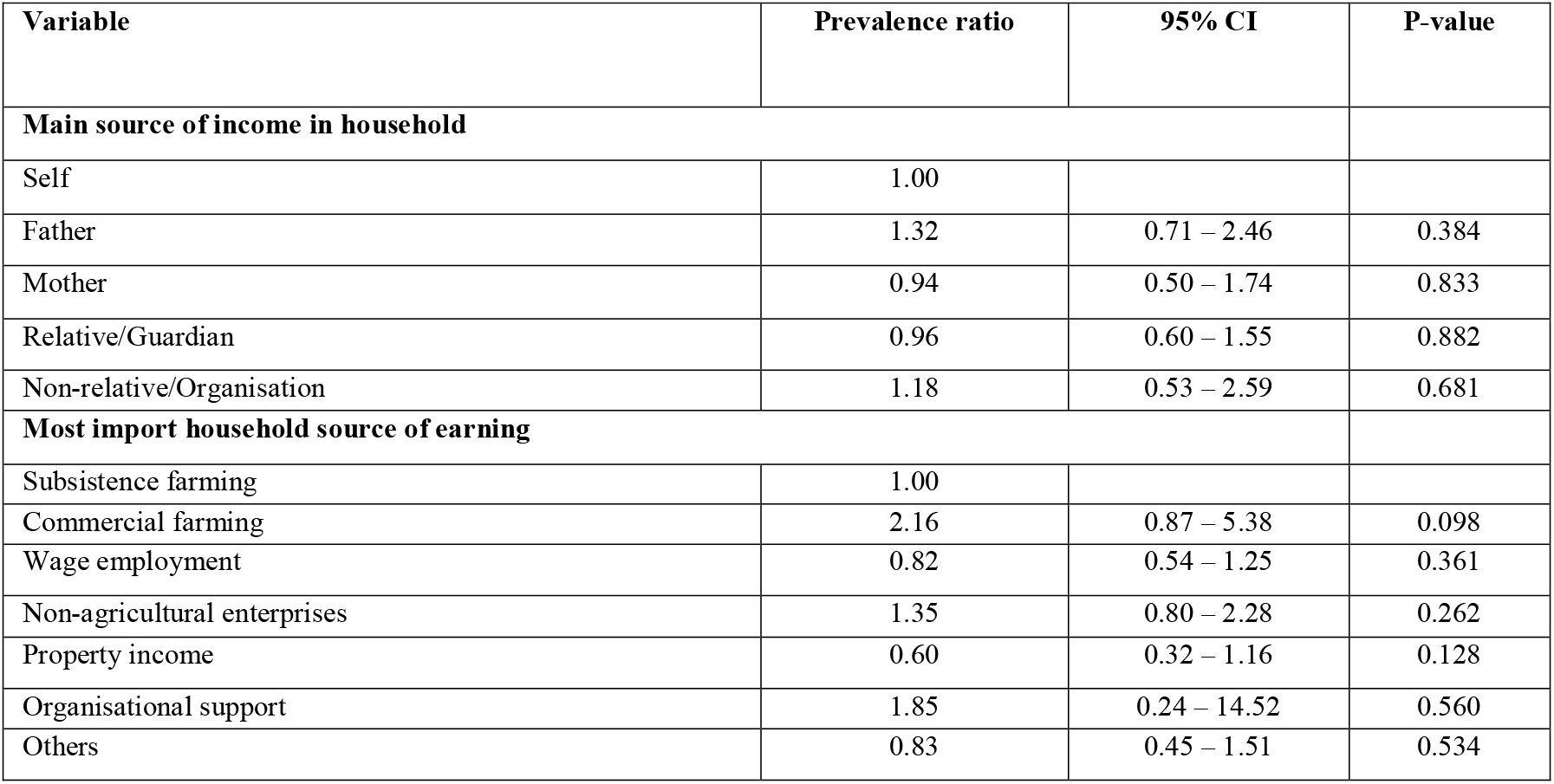
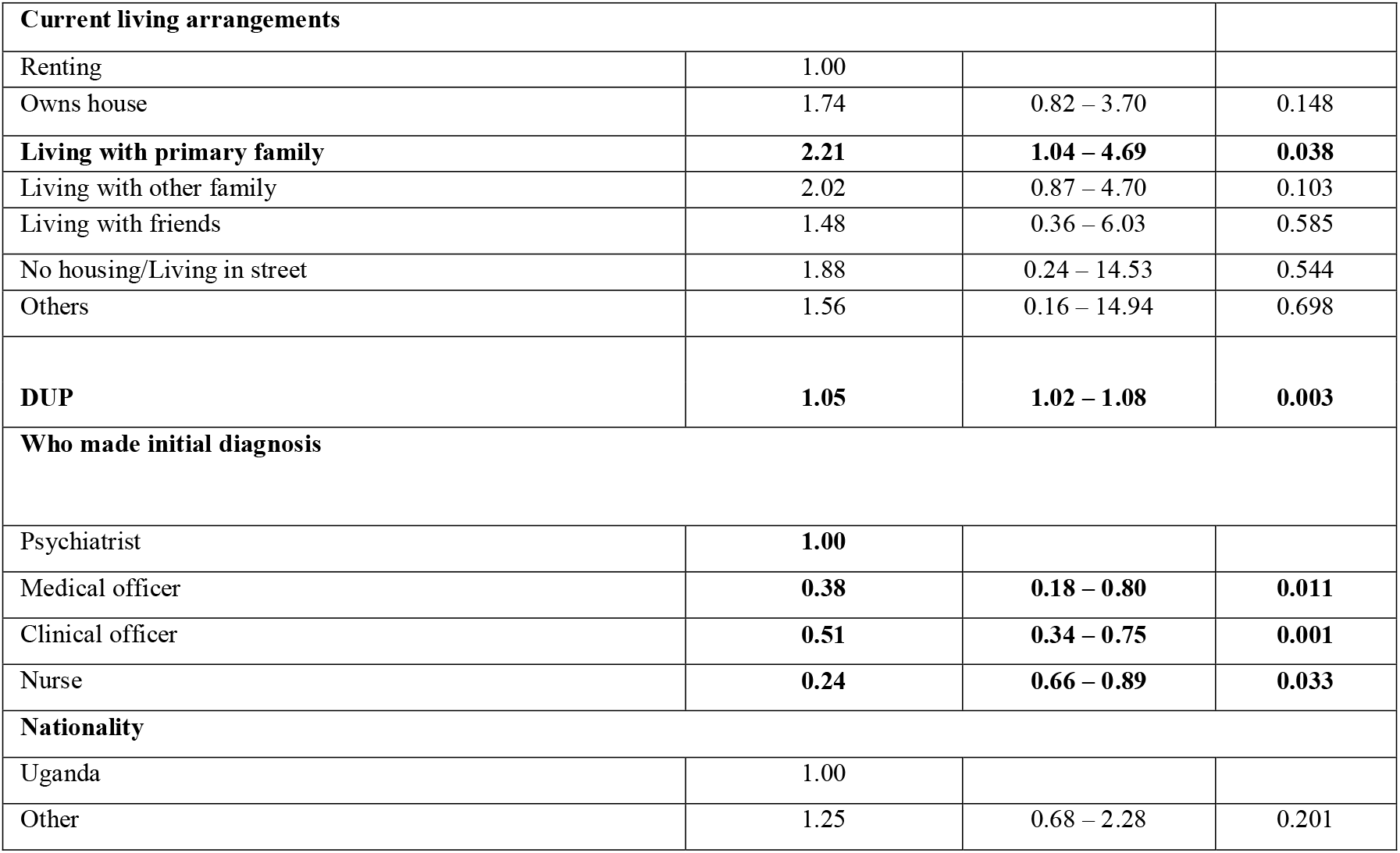
Multi-variable logistic regression between correct diagnosis and socio-demographic characteristics.

| Variable | Prevalence ratio | 95% CI | P-value |
| --- | --- | --- | --- |
| **Main source of income in household** |  |  |  |
| Self | 1.00 |  |  |
| Father | 1.32 | 0.71 – 2.46 | 0.384 |
| Mother | 0.94 | 0.50 – 1.74 | 0.833 |
| Relative/Guardian | 0.96 | 0.60 – 1.55 | 0.882 |
| Non-relative/Organisation | 1.18 | 0.53 – 2.59 | 0.681 |
| **Most import household source of earning** |  |  |  |
| Subsistence farming | 1.00 |  |  |
| Commercial farming | 2.16 | 0.87 – 5.38 | 0.098 |
| Wage employment | 0.82 | 0.54 – 1.25 | 0.361 |
| Non-agricultural enterprises | 1.35 | 0.80 – 2.28 | 0.262 |
| Property income | 0.60 | 0.32 – 1.16 | 0.128 |
| Organisational support | 1.85 | 0.24 – 14.52 | 0.560 |
| Others | 0.83 | 0.45 – 1.51 | 0.534 |
| **Current living arrangements** |  |  |  |
| Renting | 1.00 |  |  |
| Owns house | 1.74 | 0.82 – 3.70 | 0.148 |
| **Living with primary family** | **2.21** | **1.04 – 4.69** | **0.038** |
| Living with other family | 2.02 | 0.87 – 4.70 | 0.103 |
| Living with friends | 1.48 | 0.36 – 6.03 | 0.585 |
| No housing/Living in street | 1.88 | 0.24 – 14.53 | 0.544 |
| Others | 1.56 | 0.16 – 14.94 | 0.698 |
| **DUP** | **1.05** | **1.02 – 1.08** | **0.003** |
| **Who made initial diagnosis** |  |  |  |
| Psychiatrist | **1.00** |  |  |
| Medical officer | **0.38** | **0.18 – 0.80** | **0.011** |
| Clinical officer | **0.51** | **0.34 – 0.75** | **0.001** |
| Nurse | **0.24** | **0.66 – 0.89** | **0.033** |
| **Nationality** |  |  |  |
| Uganda | 1.00 |  |  |
| Other | 1.25 | 0.68 – 2.28 | 0.201 |

## DISCUSSION

### Concordance of diagnoses at admission and on the MINI

About 50% of all diagnoses made at admission by health care workers agreed with the diagnoses confirmed using an extended version of the MINI. The low levels of agreement may be due to the instability of psychotic symptoms at FEP (Björkenstam, Björkenstam, Hjern, Reutfors, & Bodén, 2013; Kim et al., 2011). Bipolar disorder and Schizophrenia had the highest diagnostic accuracy which is in keeping with other studies from high income countries (Fusar-Poli et al., 2016; Hobbs, Stanbrook, & Chakraborty, 2017; Pope, Joober, & Malla, 2013). Fourteen (14) participants (7.7%) met criteria for a substance induced psychotic disorder despite attempts to exclude all participants with substance use. Given the high prevalence of substance use among patients with FEP and its association with outcomes, it is essential that clinicians at the national referral hospital are advised to actively screen for substance use disorders in those with FEP (Mazzoncini et al., 2010; Myles, Myles, & Large, 2016)

### Duration of untreated psychosis and concordance of diagnoses

A longer duration of untreated psychosis was associated with an accurate diagnosis. A diagnosis may be harder to make early in the course of psychotic disorders which are associated with many nonspecific symptoms before the onset of frank psychosis (Dutta et al., 2007; Heckers et al., 2013; Tandon, 2013). Unfortunately, longer durations of untreated psychosis have been associated with worse outcomes (Farooq, Large, Nielssen, & Waheed, 2009; Mossaheb et al., 2013; Oosthuizen et al., 2005; Penttila, Jaaskelainen, Hirvonen, Isohanni, & Miettunen, 2014). It is therefore imperative that clinicians at the national referral hospital are taught to correctly diagnose psychotic disorders early in the course of the illness.

### Living with primary family and diagnostic accuracy

Participants living with a primary family caregiver were two times more likely to have an accurate diagnosis as compared to those with other living arrangements. This is of public health importance in this setting as about half of participants were not living with a primary family member. Primary caregivers play a crucial role in the management of patients with psychotic disorders as they are an important resource of clearer history (Cairns, Reid, & Murray, 2015; McCann, Lubman, & Clark, 2011; Strand, Olin, & Tidefors, 2015). It is plausible that those who live with a primary caregiver were brought to the hospital and thus gave clearer medical history at admission. Also, family members are often not allowed to stay during the time of the admission due to space constraints and as such are not available to attend the ward round. In tools that assess the onset of psychotic symptoms like the Nottingham Onset Schedule of psychotic disorders, it is recommended that a family member who is conversant with the patient is interviewed when making an initial assessment (Singh et al., 2005). Future strategies need to be devised were a primary family member is available during the initial patient contact.

### Level of training and a correct admission diagnosis

Psychiatrists were most accurate at diagnosing FEP followed by Psychiatric clinical officers and medical officers. However, most participants received a diagnosis from psychiatric clinical officers. This is due to the limited number of psychiatrists at the national referral and the country in general (Molodynski, Cusack, & Nixon, 2017). Effective and accurate diagnosis of FEP largely depends on the clinical skills possessed by the diagnosing clinician, with ground knowledge of all the possible underlying factors (Nasrallah, 2017). There should therefore be greater emphasis put in training the clinical officers to correctly diagnose a first episode of psychosis.

## CONCLUSION

Early intervention for psychotic disorders is crucial to long term outcomes of psychotic disorders. This early intervention requires correct diagnosis to ensure initiation of appropriate management strategies. We have highlighted the poor agreement of the diagnoses of FEP at Uganda’s national referral and teaching mental hospital. There is therefore need for additional skills training in the diagnosis and management of FEP. Specific emphasis must be placed on training lower cadre staff, emphasis on the duration of untreated psychosis in diagnostic accuracy as well as obtaining collateral history from primary family caregivers.

## LIMITATIONS

As this was a cross sectional study design it is difficult to define causality. Longitudinal studies are required to determine the accuracy of diagnoses across the admission period. No standardized method was used to determine the duration of untreated psychosis and was only calculated by subtracting the age at which the patient remembers first developing symptoms from the current age. There are however ongoing studies to determine the true DUP of this cohort using a standardized instrument (E. K. Mwesiga et al., 2019).

## Data Availability

The fully anonymized data set is available on request from the corresponding author.

## ACKNOWLEDGEMENTS

We are indebted to Prof. David Sheehan who provided us with an extended version of the MINI to carry out this study.

## DECLARATIONS

The authors declare no conflict of interests

## DATA AVAILABILITY STATEMENT

The fully anonymized data set is available on request from the corresponding author.

